# COVID-19 pandemic and lockdown measures impact on mental health among the general population in Italy. An N=18147 web-based survey

**DOI:** 10.1101/2020.04.09.20057802

**Authors:** Rodolfo Rossi, Valentina Socci, Dalila Talevi, Sonia Mensi, Cinzia Niolu, Francesca Pacitti, Antinisca Di Marco, Alessandro Rossi, Alberto Siracusano, Giorgio Di Lorenzo

## Abstract

**Background:** The psychological impact of the COronaVIrus Disease 2019 (COVID-19) outbreak and lockdown measures on the Italian population are unknown.

The current study assesses rates of mental health outcomes in the Italian general population three to four weeks into lockdown measures and explores the impact of COVID-19 related potential risk factors.

**Methods:** A web-based survey spread throughout the internet between March 27^th^ and April 6^th^ 2020. 18147 individuals completed the questionnaire, 79.6% women.

Selected outcomes were post-traumatic stress symptoms (PTSS), depression, anxiety, insomnia, perceived stress and adjustment disorder symptoms (ADS). Seemingly unrelated logistic regression analysis was performed to identify COVID-19 related risk factors.

**Results:** Respondents endorsing PTSS, depression, anxiety, insomnia, high perceived stress and adjustment disorder were 6604 (37%), 3084 (17.3%), 3700 (20.8%), 1301 (7.3%), 3895 (21.8%) and 4092 (22.9%), respectively. Being woman and younger age were associated with all of the selected outcomes. Quarantine was associated with PTSS, anxiety and ADS. Any recent COVID-related stressful life event was associated with all the selected outcomes. Discontinued working activity due to the COVID-19 was associated with all the selected outcomes, except for ADS; working more than usual was associated with PTSS, Perceived stress and ADS. Having a loved one deceased by COVID-19 was associated with PTSS, depression, perceived stress and insomnia.

**Conclusion:** We found high rates of negative mental health outcomes in the Italian general population three weeks into the COVID-19 lockdown measures and different COVID-19 related risk factors. These findings warrant further monitoring on the Italian population’s mental health.

## Background

The psychological impact of the COronaVIrus Disease 2019 (COVID-19) outbreak and related lockdown measures among the Italian population are unknown. The COVID-19 pandemic is a global health emergency that could potentially have a serious impact on public health, including mental health (World Health Organization, 2020a; Xiang et al., 2020). Since clusters of atypical pneumonia of unknown etiology were discovered in the city of Whuan, Hubei province, in late December 2019, the viral disease has continued to exponentially spread throughout China and worldwide. Italy has been the first European country that had to face the pandemic. On March 9^th^ 2020, lockdown measures were enforced by the government on entire national territory. Lockdown measures included travel restrictions, the mandatory closure of schools, nonessential commercial activities and industries. People were asked to stay at home and socially isolate themselves to prevent being infected.

As previously reported, health emergencies such as epidemic can lead to detrimental and long-lasting psychosocial consequences, due to disease related fear and anxiety, large-scale social isolation, and the overabundance of (mis)information on social media and elsewhere (Dong and Bouey, 2020). At the individual level, epidemics are associated with a wide range of psychiatric comorbidities including anxiety, panic, depression and trauma-related disorders (Tucci et al., 2017). The psychosocial impact of health emergences seems to be even higher during quarantine measures (Brooks et al., 2020). Quarantine has been associated with high stress levels (DiGiovanni et al., 2004), depression (Hawryluck et al., 2004), irritability and insomnia (Lee et al., 2005). Furthermore, being quarantined is associated with acute stress (Bai et al., 2004) and trauma-related (Wu et al., 2009) disorders, particularly in specific at-risk populations such as health workers (Lai et al., 2020).

Concerning the COVID-19 pandemic, a study on 1210 respondents in China found rates of 30% of anxiety and 17% of depression (Wang et al., 2020). Further, in a nationwide survey including more than 50.000 Chinese respondents, almost 35% of the participants reported trauma-related distress symptoms, with women and young adults showing significantly higher psychological distress (Qiu et al., 2020).

Together, these findings strongly suggest the need to accurately and timely assess the magnitude of mental health outcomes in the general population exposed to COVID-19 pandemic, with particular regard to the implementation of preventive and early interventions strategies for those at higher risk. However, no study to date has investigated mental health outcomes and associated risk factors in the Italian population. This could be of additional relevance considering the implementation of the strict lockdown and social distancing measures imposed on the entire national territory.

The aim of the current study was to assess rates of mental health outcomes in the Italian general population three to four weeks into lockdown measures and to explore the impact of COVID-19 related potential risk factors. This study aims at providing evidence that could potentially inform subsequent research strategies and mental health delivery in Italy and Europe.

## Methods

### Study Design

A cross-sectional web-based survey design was adopted. Approval for this study was obtained from the local IRB at University of L’Aquila. On-line consent was obtained from the participants.

Participants were allowed to terminate the survey at any time they desired. The survey was anonymous, and confidentiality of information was assured.

Data on mental health were collected between March 27^th^ and April 6^th^ 2020 using an on-line questionnaire spread throughout the internet, using sponsored social network advertisement together with a snowball recruiting technique. The investigated timeframe corresponds to the contagion peak in Italy, according to epidemiogical data confirmed by the World Health Organization (World Health Organization, 2020). The survey was developed using the free software Google Forms^®^.

### Participants

All Italian citizens ≥ 18 years were eligible. A total of 18147 individuals completed the questionnaire, of which 14447 (79.6%) women, median age was 38 (IQR=23). Because of the web-based design, no response rate could be estimated as it was not possible to estimate how many persons were reached by social network advertisement.

### Mental health outcomes

Post-Traumatic Stress Symptoms (PTSS), depression, anxiety, insomnia, perceived stress and adjustment disorder symptoms (ADS) were assessed using the Italian versions of the following instruments and cut-offs or scoring:

- the Global Psychotrauma Screen, post-traumatic stress symptoms subscale (GPS-PTSS) (Olff et al. *in press*): PTSS were considered of clinical relevance if more than 3 out of five 5 symptoms were reported as present;
- the 9-item Patient Health Questionnaire (PHQ-9) (Spitzer et al., 1999), using the cut-off for severe depression at ≥15;
- the 7-item Generalized Anxiety Disorder scale (GAD-7) (Spitzer et al., 2006), using the cut-off for severe anxiety at ≥15;
- the 7-item Insomnia Severity Index (ISI) (Morin et al., 2011), using the cut-off at ≥22 for severe insomnia;
- the 10-item Perceived Stress Scale (PSS) (Cohen and Hoberman, 1983), using a quartile split to separate the higher quartile from the remaining participants;
- the International Adjustment Disorder Questionnaire (IADQ) (Shevlin et al., 2020), using the standard scoring system. IADQ comprises a brief checklist of potentially stressful events, such as financial, work, health or housing problems. The IADQ checklist was modified in order to ascertain if the reported problem was due to COVID-19. ADS were rated as present if a stressful life event correlated to COVID-19 was present, together with preoccupation and failure to adapt symptoms and a relevant impact on global functioning.

### Independent variables

Standardized age, gender and region of residence (Northern Italy: Aosta Valley, Piedmont, Liguria, Lombardy, Trentino-Alto Adige, Veneto, Friuli-Venezia Giulia, Emilia-Romagna; Central Italy: Tuscany, Umbria, Marche, Lazio; Southern Italy: Abruzzo, Molise, Apulia, Campania, Basilicata, Calabria, Sicily and Sardinia) were inserted as independent variables.

Region of residence was inserted in order to account for the different incidence of COVID-19 among Italian regions. COVID-19 related independent variables were: 1) being under quarantine either because infected or in close proximity to infected people; 2) any changes in working activity compared to “working as usual” (e.g., smart-working, working activity discontinued due to lockdown measures, higher workload due to COVID-19); 3) having a loved one infected, hospitalized or deceased due to COVID-19; 4) any stressful events comprised in the IDAQ checklist, purposely modified in order to capture only stressful events due to COVID-19. The IADQ checklist comprises 8 questions about any potential stressful life event occurred in the recent past, with a yes/no response, including financial, working, educational, housing, relationship, own or loved one’ s health and caregiving problems. In order to separate COVID-19 related stressful life events from non-COVID-19 related events, responses to the checklist were modified as follows: “no”; “yes”; “yes, due to COVID-19”. Responses were collapsed in a binary variable where 1=“any stressful life evet only if due to COVID-19” and 0=“no stressful life events or presence of a stressful life event not due to COVID-19”.

### Confounders

A history of childhood trauma and any previous mental illness, as assessed by the dedicated GPS module; education level, occupation (employed, unemployed, student, retired) and being in a relationship.

### Statistical Analysis

Frequency analysis were performed in order to ascertain the prevalence of each outcome, separately for Northern, Central and Southern Italy.

A seemingly-unrelated multivariate logistic regression model was fitted in order to explore the impact of the proposed covariates and confounders on the selected outcomes. Seemingly unrelated regression models are systems of equations that allow to jointly model several outcomes, assuming correlation among their errors. Because of the very low missing data rates (<3%), missing data were treated with listwise deletion in regression analysis.

Data analysis was performed using Stata v. 16^®^ (StataCorp). Seemingly unrelated logistic regression was performed using the `-suest-` postestimation command after running a panel of logistic regressions.

## Results

Socio-demographic characteristics of the sample, along with rates of mental health outcomes, are reported in Table 1. Of the 18147 respondents, 6666 (37.14%) reported ≥3/5 PTSS, with a median total GPS symptom score of 7 (IQR=6, range 0-17); 3099 respondents (17.3%) reported severe depressive symptoms, with a PHQ total median score of 8 (IQR=6, range 0-17); 3732 (20.8%) respondents reported severe anxiety symptoms, with GAD median score of 8 (range 0-21, IQR=10); 1306 (7.3%) respondents reported severe insomnia symptoms, with ISI median total score of 10 (range 0-28, IQR=12); PSS total score median was 25 (range 4-44, IQR=13), 75^th^ percentile was 31, with 3933 (21.9%) respondents scoring above this threshold; 4129 (23.0%) respondents reported a IADQ scoring compatible with the suspect of a presence of an adjustment disorder.

**Table 1.**
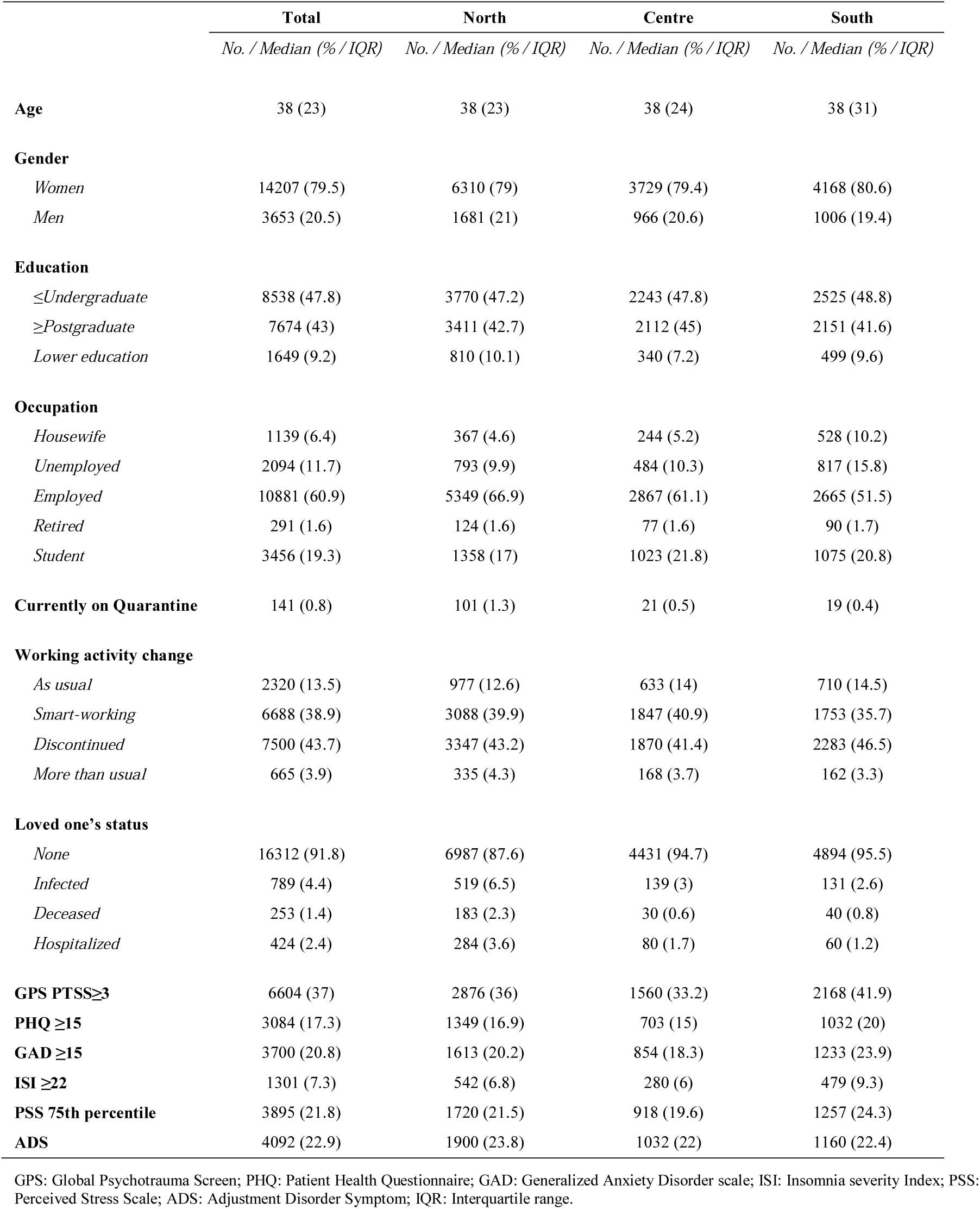
Demographic characteristics and rates of mental health outcomes in the sample.

Seemingly unrelated logistic regression analyses are reported in Table 2. Being a woman was associated with all of the selected outcomes (PTSS: OR=2.12 [1.94, 2.31]; depression: OR=1.39 [1.24, 1.56]; anxiety: OR=1.77 [1.59, 1.97]; perceived stress: OR=2.06 [1.85, 2.30]; insomnia: OR=1.50 [1.26, 1.78]; adjustment disorder: OR=1.64 [1.45, 1.84]). Younger age was associated with PTSS, depression, anxiety and perceived stress (respectively: OR=1.49 [1.39, 1.60]; 1.55 [1.42, 1.69]; 1.72 [1.59, 1.87]; 1.76 [1.62, 1.90]). Compared to Northern Italy, participants from Southern Italy showed higher odds of all of the selected outcomes, except for ADS (PTSS: OR=1.36 [1.26, 1.47]; depression: OR=1.25 [1.13, 1.37]; anxiety: OR=1.29 [1.18, 1.41]; perceived stress: OR=1.20 [1.10, 1.32]; insomnia: OR=1.41 [1.24, 1.62]). Being under quarantine because infected or in close proximity to infected people was associated with PTSS, Anxiety and ADS (respectively: OR=1.74 [1.21,2.49]; 1.52 [1.05,2.22]; 2.28 [1.44,3.61]). Having experienced a stressful life event due to COVID-19, as assessed by the modified IADQ checklist, was associated with all of the selected outcomes (PTSS: OR=1.46 [1.37,1.56]; depression: OR=1.58 [1.45,1.72]; anxiety: OR=1.64 [1.51,1.78]; perceived stress: OR=1.82 [1.68,1.97]; insomnia: OR=1.58 [1.40,1.79]). OR of IADQ-Checklist on ADS was not estimated due to the perfect prediction, because having an IADQ checklist event is a prerequisite for having a suspected Adjustment Disorder. Working activity discontinued due to COVID-19 was associated with all of the selected outcomes except for ADS (PTSS: OR=1.15 [1.05,1.27]; depression: OR=1.40 [1.23,1.59]; anxiety: OR=1.16 [1.03,1.31]; perceived stress: OR=1.19 [1.06,1.34]; insomnia: OR=1.22 [1.03,1.46]), while working more than usual due to the COVID-19 was associated with PTSS, perceived stress and ADS (respectively: OR= 1.42 [1.18,1.71]; 1.71 [1.38,2.12]; 1.39 [1.04,1.87]). Having a loved one deceased by COVID-19 was associated with PTSS (OR=1.68 [1.30,2.16]), depression (OR=1.41 [1.03,1.93]), perceived stress (OR=1.34 [1.01, 1.78], insomnia (OR=1.74 [1.18, 2.54]), while having a loved one diagnosed with COVID-19 was associated with PTSS (OR=1.22 [1.05, 1.42]).

**Table 2:** Seemingly Unrelated Logistic Regression.

|  | PTSS<br>*OR [95%CI]* | Depression<br>*OR [95%CI]* | Anxiety<br>*OR [95%CI]* | Perceived Stress<br>*OR [95%CI]* | Insomnia<br>*OR [95%CI]* | ADS<br>*OR [95%CI]* |
| --- | --- | --- | --- | --- | --- | --- |
| **Age<sup>§</sup>** | 1.49*** [1.39,1.60] | 1.55*** [1.42,1.69] | 1.72*** [1.59,1.87] | 1.76*** [1.62,1.90] | 1.01 [0.97,1.05] | 1.05 [0.75,1.47] |
| **Gender** |  |  |  |  |  |  |
| *Men* | 1.00 (ref) |  |  |  |  |  |
| *Women* | 2.12*** [1.94,2.31] | 1.39*** [1.24,1.56] | 1.77*** [1.59,1.97] | 2.06*** [1.85,2.30] | 1.50*** [1.26,1.78] | 1.64*** [1.45,1.84] |
| **Region** |  |  |  |  |  |  |
| *North* | 1.00 (ref) |  |  |  |  |  |
| *Centre* | 0.93 [0.86,1.01] | 0.87* [0.78,0.97] | 0.90* [0.82,1.00] | 0.90* [0.82,0.99] | 0.9 [0.77,1.05] | 0.91 [0.81,1.02] |
| *South* | 1.36*** [1.26,1.47] | 1.25*** [1.13,1.37] | 1.29*** [1.18,1.41] | 1.20*** [1.10,1.32] | 1.41*** [1.24,1.62] | 0.95 [0.85,1.06] |
| **COVID-19-Related Stressful Event** | 1.46*** [1.37,1.56] | 1.58*** [1.45,1.72] | 1.64*** [1.51,1.78] | 1.82*** [1.68,1.97] | 1.58*** [1.40,1.79] | n.a. n.a. |
| **Currently On Quarantine** | 1.74** [1.21,2.49] | 1.49 [0.98,2.26] | 1.52* [1.05,2.22] | 1.42 [0.97,2.07] | 1.23 [0.69,2.18] | 2.28*** [1.44,3.61] |
| **Working Activity Change** |  |  |  |  |  |  |
| *As Usual* | 1.00 (ref) |  |  |  |  |  |
| *Smart-Working* | 1.01 [0.91,1.12] | 0.99 [0.86,1.14] | 0.97 [0.85,1.10] | 1.02 [0.90,1.15] | 0.9 [0.74,1.10] | 1.07 [0.91,1.25] |
| *Discontinued* | 1.15** [1.05,1.27] | 1.40*** [1.23,1.59] | 1.16* [1.03,1.31] | 1.19** [1.06,1.34] | 1.22* [1.03,1.46] | 1.1 [0.95,1.28] |
| *More Than Usual* | 1.42*** [1.18,1.71] | 1.26 [0.98,1.63] | 1.25 [1.00,1.57] | 1.71*** [1.38,2.12] | 1.29 [0.93,1.80] | 1.39* [1.04,1.87] |
| **Loved One's Condition** |  |  |  |  |  |  |
| *None* | 1.00 (ref) |  |  |  |  |  |
| *Infected* | 1.22* [1.05,1.42] | 1.05 [0.87,1.28] | 0.91 [0.75,1.10] | 0.88 [0.73,1.05] | 1.02 [0.77,1.35] | 0.96 [0.79,1.17] |
| *Deceased* | 1.68*** [1.30,2.16] | 1.41* [1.03,1.93] | 1.22 [0.91,1.65] | 1.34* [1.01,1.78] | 1.74** [1.18,2.54] | 1.21 [0.87,1.68] |
| *Hospitalized* | 1.22 [1.00,1.48] | 1.09 [0.84,1.41] | 1.25 [0.99,1.57] | 1.1 [0.87,1.39] | 1.1 [0.76,1.60] | 1.16 [0.91,1.49] |
| **In A Relationship** | 1.14*** [1.06,1.22] | 0.92 [0.84,1.00] | 1.11* [1.02,1.22] | 1.11* [1.02,1.21] | 1.08 [0.94,1.23] | 1.07 [0.97,1.19] |
| **Education** |  |  |  |  |  |  |
| *≥Postgraduate* | 1.00 (ref) |  |  |  |  |  |
| *≤Undergraduate* | 1.12** [1.04,1.20] | 1.30*** [1.19,1.43] | 1.28*** [1.18,1.39] | 1.25*** [1.15,1.36] | 1.31*** [1.15,1.50] | 1.05 [0.95,1.16] |
| *Lower Education* | 1.25*** [1.11,1.41] | 1.62*** [1.40,1.87] | 1.51*** [1.32,1.74] | 1.47*** [1.28,1.69] | 1.76*** [1.46,2.13] | 1.21* [1.01,1.44] |
| **Occupation** |  |  |  |  |  |  |
| *Employed* | 1.00 (ref) |  |  |  |  |  |
| *Housewife* | 1.28*** [1.11,1.47] | 1.35** [1.12,1.63] | 1.31** [1.11,1.55] | 1.21* [1.03,1.44] | 1.39** [1.11,1.74] | 1.05 [0.83,1.32] |
| *Unemployed* | 1.05 [0.94,1.17] | 1.59*** [1.40,1.80] | 1.39*** [1.23,1.57] | 1.22** [1.08,1.37] | 1.33** [1.12,1.58] | 1.09 [0.93,1.27] |
| *Retired* | 0.9 [0.66,1.22] | 1.17 [0.79,1.75] | 1.02 [0.69,1.51] | 1.39 [0.96,2.01] | 0.88 [0.52,1.48] | 0.46* [0.22,0.97] |
| *Student* | 0.79*** [0.71,0.88] | 1.60*** [1.41,1.83] | 1.02 [0.90,1.16] | 1.28*** [1.13,1.44] | 1.02 [0.86,1.22] | 1.16 [0.84,1.62] |
| **Childhood Trauma** | 1.06 [0.99,1.13] | 1.41*** [1.30,1.54] | 1.29*** [1.19,1.39] | 1.01 [0.93,1.09] | 1.50*** [1.33,1.70] | 1.10* [1.01,1.21] |
| **Prior Psychiatric Diagnosis** | 1.59*** [1.48,1.71] | 2.19*** [2.01,2.39] | 2.10*** [1.94,2.28] | 1.73*** [1.59,1.87] | 1.76*** [1.56,1.98] | 1.25*** [1.13,1.39] |

\*p<0.05; \*\*p<0.005; \*\*\*p<0.001; PTSS: Post-Traumatic Stress Symptoms; ADS: Adjustment Disorder Symptom; § Age is standardized and reversed, younger age has an OR>1 if associated with heightened risk.

## Discussion

In this study, we report for the first time on the mental health outcomes related to COVID-19 outbreak and related lockdown measures on the general population in Italy. To the best of our knowledge, this is the first study to report on mental health outcomes related to the COVID-19 outbreak in Europe on such a large sample size. This study shows relatively high rates of PTSS, Depression, Anxiety, Insomnia, Perceived stress and ADS, with young women having higher odds of endorsing a mental health outcome. These outcomes were associated with a number of COVID-19-related risk factors, including being under quarantine, having a loved one deceased by COVID-19, working activity discontinued due to lockdown measures, or experiencing other stressful events (i.e. working, financial, relationship or housing problems) due to the pandemic or lockdown measures. These findings were adjusted for previous psychiatric illness and a history of childhood trauma, suggesting that the COVID-19 pandemic is exerting an independent effect on the population mental health.

### Previous literature

Compared to an early report on the mental health outcomes related to COVID-19 in China on 1210 respondents (Wang et al., 2020), we found lower rates of anxiety, similar rates of depression and higher levels of perceived stress, notwithstanding differences in assessment tools. The negative association with age and the positive association with female gender was confirmed, suggesting that young women may be at heightened risk for mental disorders. Compared to another large web-based survey from China on 52730 respondents that evaluated peritraumatic stress-related symptoms, we found similar rates of PTSS (Qiu et al., 2020). Another study on 285 participants from hardest-hit Hubei province found substantially lower rates of PTSS, around 7% (Liu et al., 2020). Such disparities could be due to different assessment tools used and differences in sample size. A study on 7143 medical students in China (Cao et al., 2020) found severe anxiety rates, assessed as GAD≥15, to be 0.9%, compared to our 20.9%. This inconsistence could be due to the particular population investigated, having a high education level. Indeed, higher education was associated with better outcomes in our study. Furthermore, cultural, social and health care system differences between China and Italy could explain differences in reported mental health outcomes.

Coherently with previous reports from China, female gender (Liu et al., 2020; Qiu et al., 2020; Wang et al., 2020) and younger age (Qiu et al., 2020; Wang et al., 2020) were consistently associated with higher risk for different mental health outcomes. If confirmed in other populations worldwide, these findings could be of great importance for subsequent intervention strategy for global mental health related to COVID-19.

### Relevance

Monitoring populations’ mental health is critical during a pandemic, as generalized fear and fear-induced over-reactive behaviour among the public could impede infection control (Dong and Bouey, 2020). Further, the current strict lockdown measures and the home confinement of unknown duration represent an unprecedented stressful event potentially leading to significant long-term health costs. Epidemiological monitoring and targeted intervention should be therefore timely implemented to prevent further mental health problems. Indeed, once the outbreak will be over, its negative socio-economic consequences may have a detrimental effect on the population’s mental health, as suggested by our finding of an heightened risk of mental health issues due to COVID-19 related working difficulties and by earlier studies related to the last economic crisis (Wahlbeck et al., 2011).

### Limitations and future directions

This study has some important limitations due to the sampling technique. Relying on social networks voluntary recruitment and re-sharing could have introduced an important selection bias, firstly excluding people not on social networks, and secondly introducing a self-selection bias, as suggested by the highly unbalanced gender ratio observed. This latter bias could have affected also two other large web-based surveys in China, that reported on samples with a 64.7% and 67.3% proportion of woman (Qiu et al., 2020; Wang et al., 2020). For these reasons, rates of mental health outcomes should be interpreted with caution. Secondly, this survey was based on self-report instruments that could introduce a systematic bias and return different rates compared to interview-based measures.

This study has also several strengths, including a very large sample size and the sampling timeframe that corresponded to the pandemic peak in Italy.

Future studies will need to monitor the trajectory of mental health outcomes, in order to define mental health interventions at a population level.

## Conclusions

We found high rates of negative mental health outcomes in the Italian general population three to four weeks into the COVID-19 pandemic and lockdown measures. COVID-19 related factors were associated with these outcomes independently from previous mental illness or childhood trauma. These findings warrant further monitoring on the Italian population’s mental health and could serve to inform structured interventions in order to mitigate the impact on mental health of the outbreak.

## Data Availability

Data available on request from the authors.

## Authors contribution

Conceptualization: RR, VS, FP, GDL; Methodology: RR; Formal Analysis: RR; Data Curation: RR, SM, GDL; Writing - Original Draft: RR, VS; Writing - Review & Editing: RR, VS, DT, ADM, FP, SM, CN, AR, AS, GDL.

## Acknowledgments

This work is supported by Territori Aperti, a project founded by “Fondo Territori Lavoro e Conoscenza CGIL CISL UIL”.

## Contribution to the field

The COronaVIrus Disease 2019 (COVID-19) pandemic is a global health emergency that could potentially have a serious impact on public health, including mental health. The psychological impact of the COVID-19 outbreak and related lockdown measures among the Italian population are unknown. In this web-based study, we report for the first time on the psychological impact of COVID-19 outbreak on the general population in Italy. This study shows high rates of post-traumatic symptoms, Depression, Anxiety, Insomnia, Perceived stress and Adjustment Disorder associated with a number of COVID-19-related risk factors. This study represents the first European report on mental health in the time of the COVID-19, and it could have a strong impact on subsequent research and clinical intervention strategy for global mental health related to COVID-19.

